# Comparison of IFSG to SOC for treatment of venous leg ulcers using real-world data from the USWR with 1:1 matching on 14 wound/patient factors

**DOI:** 10.1101/2025.10.29.25339052

**Authors:** Hongyu Miao, Matthew Pine, John C. Lantis, Caroline Fife

## Abstract

**Background:** Venous leg ulcers (VLUs) impose substantial morbidity and Medicare spending, yet many real-world ulcers remain refractory to standard of care (SOC). Intact fish-skin graft (IFSG) is a biologic graft used for chronic wounds. We evaluated the comparative effectiveness of IFSG versus SOC in routine practice using a specialty wound registry with Real World Evidence design features intended to minimize bias.

**Methods:** We performed a retrospective, target-trial–emulating, 1:1 propensity score–matched comparative-effectiveness study within the U.S. Wound Registry (USWR). Matching used 14 prespecified patient- and wound-level covariates (including mobility as a measure of frailty and number of concomitant wounds).

**Results:** The matched cohort included 129 IFSG-treated VLUs and 129 SOC-treated VLUs. Baseline balance was excellent by standardized mean differences. Small residual differences favored SOC; IFSG-treated wounds were older and trended larger. Healing occurred in 85.3% of IFSG-treated wounds (110/129) versus 75.2% of SOC-treated wounds (97/129); the absolute difference (+10.1%) was just below statistical significance (p=0.0801). SOC-treated VLUs increased in size on average more than IFSG-treated VLUs (p=0.0036).

**Conclusions:** In a national wound registry with rigorous cohort construction, aligned index timing, comprehensive covariate control, and structured outcome capture, IFSG demonstrated favorable real-world effectiveness versus SOC for VLUs with a trend towards more healed wounds and a statistically significant lower average wound expansion. The high healing rate in the SOC arm is plausibly explained by baseline advantages (shorter duration, smaller area, and “never-advanced-therapy” selection) as well as the absence of a set follow-up duration that typically extended until healing, a competing event, or administrative end of observation.

## 1. Introduction

Venous leg ulcers (VLUs) affected an estimated 627,000 Medicare beneficiaries in 2019, a 25.4% increase from 2014, and accounted for $1.13 billion in Medicare spending that same year.^1^ VLUs arise from chronic venous insufficiency and subsequent venous hypertension, leading to skin and subcutaneous tissue breakdown. They often drain copiously, have a significant impact on quality of life and usually require medical intervention to heal.^2,3^ The standard of care (SOC) for VLUs, well defined by numerous guidance documents, centers on compression therapy and moist wound care.^4^ Recent guidance would indicate that early correction of identified superficial venous reflux not only reduces time to closure but also decreases recurrence.^5,6^ In VLU prospective trials difficult-to-close VLUs (usually defined as less than 20% wound are reduction at 2 weeks); heal at rates of 40–70% within 12–16 weeks and about 80% in 24 weeks.^7,8^

However, in the real world, healing rates are lower and a substantial portion of VLUs can remain unhealed for six to twelve months or longer.^9,10^ For ulcers that fail to improve after approximately 4–6 weeks of optimal SOC, most guidance documents recommend adjunctive therapies such as biologic tissue grafts,^2^ the efficacy of which has been established by numerous prospective trials.^11,12^ However, unlike prospective trial subjects, real world VLU patients often have complex comorbid diseases such as diabetes, obesity, impaired mobility, heart disease and peripheral arterial disease, conditions which would exclude them from most prospective studies.^13,14^ Since these conditions are common among Medicare beneficiaries with VLUs, there is a need to perform real-world comparative effectiveness studies to demonstrate the potential value of biologic tissue grafts in routine clinical practice among patients with highly refractory VLUs.

Real-world data (RWD) derived directly from point-of-care electronic health records offer relevant insight but come with practical limitations that can compromise credibility if unmanaged.^15^ Practical limitations of RWD in wound care often stem from inconsistent cohort construction, heterogeneous outcome definitions, mis-classification of wound and patient variables, and incomplete reporting—issues that can inflate bias and hinder cross-study comparability. In her guidelines for improving the scientific validity of observational studies, Carter recommends the following: define cohorts with minimal inclusion/exclusion criteria to pre-serve representativeness, preplan subgroup analyses, and build well-matched comparators using clearly classified wound/patient covariates. She further recommends publishing a core benchmark set (e.g., wound size, duration, infection status, compression fidelity). Adopting these practices—along with transparent SAPs and complete data dictionaries—substantially reduces misclassification and confounding, improves reproducibility, and strengthens policy fitness for Medicare coverage evaluations.^15^

Constructing credible comparative cohorts in RWD requires careful control for treatment-selection bias and confounding at both the patient and wound levels. In addition to peripheral arterial disease, wound age and infection or bioburden, prior analyses of U.S. Wound Registry (USWR) data identified two previously un-recognized factors strongly associated with delayed VLU healing: the number of concomitant wounds and mobility status, an indirect measure of patient frailty.^16^

Intact fish-skin graft (IFSG) is a biologic tissue graft from decellularized North Atlantic cod skin that preserves an intact extracellular matrix to support host-cell infiltration and tissue regeneration and is indicated for chronic wounds, including VLUs.^17^ This study evaluates the real-world comparative effectiveness of IFSG versus SOC for VLUs using USWR data which allows matching a previously unparalleled number of clinical factors. Our primary objectives were to compare the healing rate and percent area reduction between IFSG-treated and SOC-treated wounds among hard-to-heal VLUs in complex patients that have failed many weeks of advanced treatment. Here we match IFSG-treated and SOC-treated cohorts by an unprecedented 14 variables and find that IFSG-treated VLUs trend toward higher healing and have statistically significantly greater area reduction than VLUs treated with SOC, proving that IFSG exhibits effectiveness in the real world.

## Materials and Methods

### Real world data source

Data was sourced spanning the years 2006 to June 2025 from the USWR, which is a repository of deidentified patient data transferred from the electronic health record (EHR) Intellicure™ (Intellicure, Inc., The Woodlands, TX). Intellicure™ is specific to wound care and used by over 500 wound care facilities and practitioners in 29 U.S. States and Puerto Rico.^18^ This EHR not only enables the documentation of clinical information at point of care, but also offers up-to-date clinical decision support to wound care providers. Clinical wound care information recorded at point of care is electronically transferred to the USWR, creating a repository of real-world patient data. The USWR maintains a HIPAA-compliant limited dataset; data are proprietary but may be shared under data-use agreements as noted in the Data Availability statement.

### Cohort eligibility and propensity matching

We identified VLUs in the USWR using pre-specified ICD-9/ICD-10 diagnosis codes combined with Intellicure™ wound-location fields for the lower leg. All consecutive VLU encounters meeting these definitions during the study window were eligible. We did not exclude based on wound size, wound age, or patient comorbidities, to preserve representativeness of routine practice and Medicare relevance.

The exposure cohort comprised all VLUs treated with intact fish-skin graft (IFSG) (129 wounds in 94 patients). Fish skin graft treated wounds were identified by using pre-specified CTP codes, Q4158 and A2019 combined with Intellicure™ treatment-location fields for MariGen, Kerecis Omega3, and MariGen Shield.

Patients with missing treatment outcomes, due to reasons such as being lost to follow-up, deceased, or transferred, were excluded from subsequent analyses. Wound location was extracted from the ICD-10 codes associated with each wound, primarily L97.2, L97.3, L97.8 and L97.9. Wound age was calibrated from the date noted (that is, the first visit at which the VLU’s presence was recorded) to the date of the first Kerecis application. Correspondingly, baseline wound characteristics such as wound size and infection score were extracted at the visit of first IFSG application.

In accordance with guidelines based on the RECORD statement for improved scientific validity of observational studies, we matched the IFSG-treated cohort to SOC-treated patients/wounds according to the following recommended characteristics known to affect the odds of healing: sex, patient age, mobility status (defined as method of arrival to appointments), diabetes, chronic kidney disease, number of concomitant wounds (highest number of concurrent wounds noted any time after the first application of IFSG), wound area (at the time of first IFSG application), and wound age (at the time of first IFSG application).^11^ We also matched to the following additional characteristics: autoimmune disease, congestive heart failure, dementia, paralysis, wound location, and malnourishment. All matched covariates are given in Tables 1 and 3. Matching was done in a 1:1 ratio, resulting in a SOC-treated cohort of 129 wound in 129 patients.

**Table 1.**
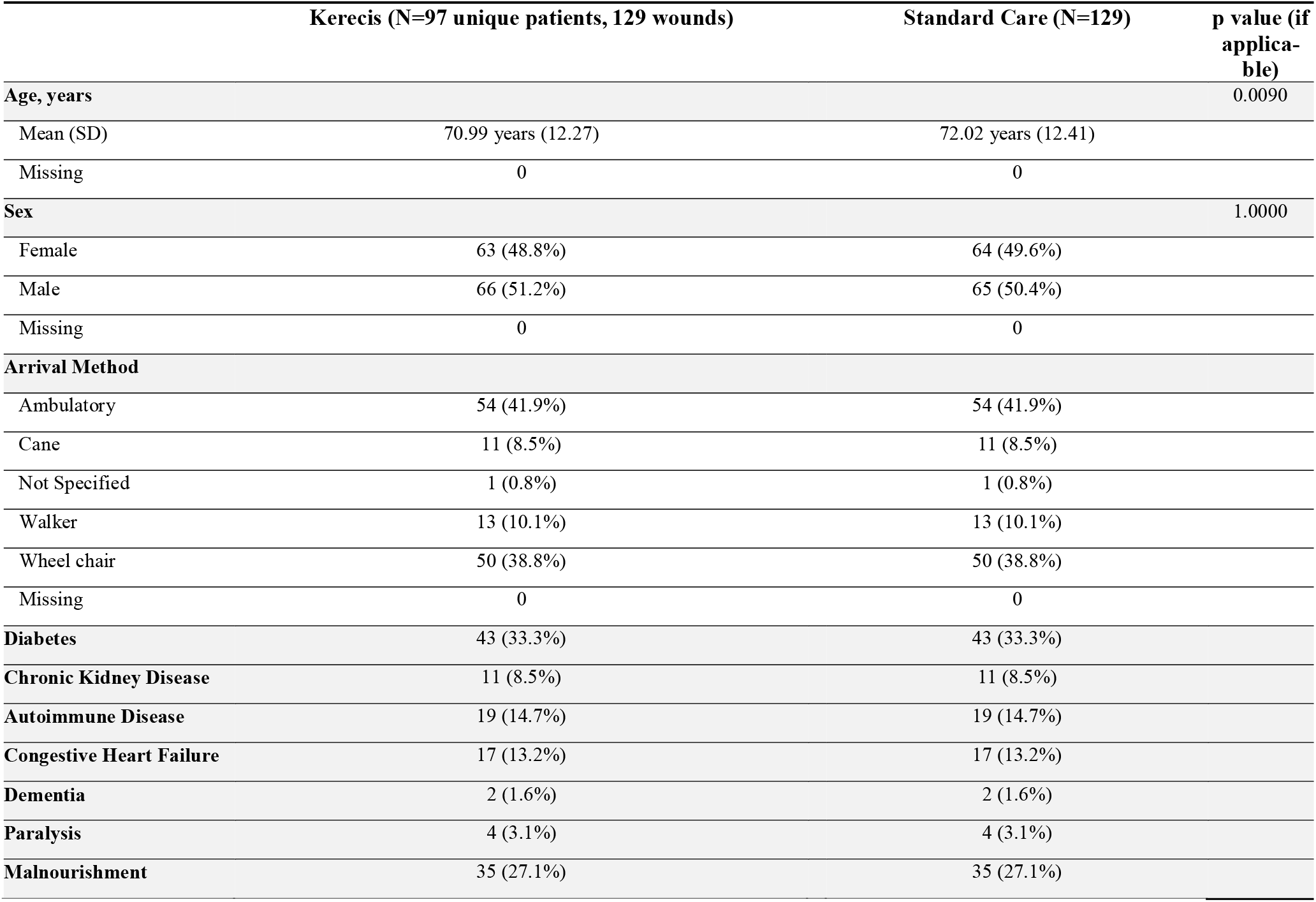
Patient characteristics used for matching cohorts.

Statistically, the 1:1 match of SOC-treated VLUs to IFSG-treated VLUs was obtained using the nearest neighbor approach based on Mahalanobis distance since propensity-score based matching was found computationally ineffective (R v4.5.1, MatchIt package v4.7.2).

### Comparison between IFSG-treated and SOC-treated VLUs

Based on the identified matched pairs, descriptive statistics were generated to summarize the patient-level and wound-level characteristics (e.g., the mean and standard deviation for continuous variables and the count and percentage for discrete variables). The signed rank test was used to account for the deviation from the normality assumption in data distribution and to compare the outcomes between Kerecis applications and standard care, including wound size changes, wound time in service and healing rate. Unless otherwise specified, the Type I error rate was controlled at 0.05.

### Index date

To eliminate immortal-time and alignment bias, we defined the index date separately by arm but anchored to the same clinical status:

IFSG arm: index = date of first IFSG application; baseline covariates (e.g., wound area/age, infection score, compression status) were taken from this visit.

SOC arm: for each matched SOC wound, index = the first SOC visit at which the wound met the same clinical status profile as its IFSG match (i.e., VLU under active SOC, with comparable wound age/area and care context), operationalized via the matched covariates and Intellicure structured fields.

### Outcome measurements

The primary outcome was complete wound healing, defined as full epithelialization with no drainage and no further need for dressings, as documented in structured EHR fields during routine clinic visits. A wound was considered healed when a wound was either indicated to have healed in the EHR or if the wound area had completely reduced and the wound did not have subsequent treatment To be conservative, only positively confirmed outcome descriptions, including “Goal Achieved”, “Healed”, and “Resolved”, were re-labeled as “Healed”; all other negative, uncertain, or ambiguous outcome descriptions (e.g., “Failed”, “Converted”, “Diagnosis Change”, “Bettering”) were relabeled as “Not Healed” in the processed data. Recurrence after documented healing was not part of the primary endpoint; once healed, wounds remained classified as healed for the primary analysis.

A secondary outcome was percent area reduction (PAR), analyzed at fixed intervals. Wound area measurements were obtained from structured EHR entries based on length-by-width measurement recorded in consistent units across visits. To address biologically implausible fluctuations in area that can arise from docu-mentation or tracing error, values triggering automated data-quality flags (for example, extreme week-to-week changes) were examined using approaches such as trimming while the primary analysis used the recorded values.

Outcome dates (healing, amputation, death) were not imputed. When area at a target time point was missing but surrounding visits existed, PAR was derived from the nearest visit within the predefined window; other-wise, PAR at that time point was treated as missing without imputing values, and the observation was excluded from that specific analysis while remaining in other analyses.

### Missing data, censoring, and follow-up

Baseline covariates with missing values were handled using multiple imputation, with imputation models including all propensity covariates, the exposure indicator, and outcomes (without using outcome timing). This approach was selected given notable missingness in several descriptive variables (e.g., Mini Nutritional Assessment and certain demographics) in the source registry tables. Complete-case and missing-indicator analyses were prespecified as sensitivity analyses to assess robustness to alternative assumptions about missingness.

Outcome dates (healing, amputation, death) were not imputed. Participants without a healing or competing event were censored at the last observed clinic visit. Transfers outside the registry network and loss to follow-up were treated as right-censoring events.

## Results

### Patient Characteristics of Cohorts

Comorbidities and other patient characteristics matched as covariates between cohorts are given in Table 1. The matched cohorts were closely comparable across all prespecified covariates used in propensity matching (Table 1). A small residual difference in age remained after matching (mean [SD]: 70.99 [12.27] years in the IFSG group vs 72.02 [12.41] years in the SOC group), amounting to just over one year on average and unlikely to be clinically meaningful. Sex distribution was nearly identical (female: 48.8% IFSG vs 49.6% SOC). Arrival method, diabetes, chronic kidney disease, autoimmune disease, congestive heart failure, dementia, paralysis, and documented malnourishment were balanced by design.

Additional demographic and wound-relevant characteristics (Table 2) indicate that the study population is predominantly Medicare-relevant: 73.6% of IFSG and 76.0% of SOC patients were aged ≥65 years. Most patients identified as White (82.5% IFSG; 84.2% SOC), with Black or African American patients comprising the next largest group (16.5% IFSG; 11.9% SOC); few patients identified as Hispanic or Latino. Mean BMI was 31.92 in the IFSG cohort and 34.43 in the SOC cohort; this difference did not reach statistical significance and was not associated with imbalance in other nutrition-related measures. Mini Nutritional Assessment scores were similar between groups (means of 11.67 and 12.61 for the IFSG-treated cohort and the SOC-treated cohort, respectively). Both cohorts included current smokers and a small number of patients reporting illicit drug use. Proportions of people with depression and with obesity were the same in each group. Overall, baseline characteristics demonstrate successful matching with only trivial residual differences and strong overlap between groups, supporting the validity of subsequent comparative effectiveness analyses.

**Table 2.** Additional patient characteristics.

|  | Kerecis (N=97 unique patients, 129 wounds) | Standard Care (N=129) | p value |
| --- | --- | --- | --- |
| **Patients ≥65** | 95 (73.6%) | 98 (76.0%) | 0.4497 |
| Missing | 0 | 0 |  |
| **Race** |  |  |  |
| Asian | 0 (0.0%) | 1 (1.0%) |  |
| Black or African American | 17 (16.5%) | 12 (11.9%) |  |
| Other | 1 (1.0%) | 3 (3.0%) |  |
| White | 85 (82.5%) | 85 (84.2%) |  |
| Missing | 26 | 28 |  |
| **Ethnicity** |  |  |  |
| Declined to Specify | 2 (2.3%) | 0 (0.0%) |  |
| Hispanic or Latino | 3 (3.5%) | 1 (1.3%) |  |
| Not Hispanic or Latino | 81 (94.2%) | 78 (98.7%) |  |
| Missing | 43 | 50 |  |
| **BMI** |  |  |  |
| Mean (SD) | 31.92 (8.77) | 34.43 (10.08) | 0.0591 |
| Missing | 1 | 1 |  |
| **MNA Score** |  |  |  |
| Mean (SD) | 11.67 (2.19) | 12.61 (1.73) | 0.2055 |
| Missing | 34 | 78 |  |
| **Tobacco Use** | 16 (12.4%) | 10 (7.8%) | 0.2864 |
| **Peripheral Artery Disease** | 27 (20.9%) | 27 (20.9%) | 1.0000 |
| **Illicit Drug Use** | 2 (1.6%) | 1 (0.8%) | 1.0000 |
| **Depression** | 10 (7.8%) | 10 (7.8%) | 1.0000 |
| **Obesity** | 68 (52.7%) | 68 (52.7%) | 1.0000 |

### Wound Characteristics of Cohorts

Wound clinical and baseline characteristics matched as covariates between cohorts are given in Table 3. Wound location was almost exactly matched between cohorts. IFSG-treated wounds were much older on average than SOC-treated wounds, a difference that was statistically significant. IFSG-treated wounds were also larger in surface area than SOC-treated wounds, although this difference was not statistically significant. The distribution of number of concomitant wounds within a patient was similar between IFSG-treated patients and SOC-treated wounds. Notably, less than a quarter of VLUs occurred in isolation; the vast majority of patients had more than one wound. Additional wound clinical and baseline characteristics compared between cohorts are given in Table 4.

**Table 3.** Wound baseline and clinical characteristics used for matching cohorts.

|  | Kerecis (N=129) | Standard Care (N=129) | p value |
| --- | --- | --- | --- |
| **Wound Location** |  |  |  |
| ANKLE | 27 (20.9%) | 27 (20.9%) |  |
| CALF | 15 (11.6%) | 14 (10.9%) |  |
| HEEL AND MIDFOOT | 6 (4.7%) | 6 (4.7%) |  |
| OTHER PART OF LOWER LEG | 81 (62.8%) | 82 (63.6%) |  |
| Missing | 0 | 0 |  |
| **Wound Age at First Application** |  |  |  |
| Mean (SD) | 117.44 days (141.05) | 88.97 days (110.16) | < 1e-04 |
| Missing | 0 | 0 |  |
| **Wound Size at First Application** |  |  |  |
| Mean (SD) | 14.22 cm (37.66) | 11.10 cm (32.39) | 0.1289 |
| Missing | 0 | 0 |  |
| **Total Concomitant Wounds** |  |  |  |
| 1 | 31 (24.0%) | 31 (24.0%) |  |
| 2 | 31 (24.0%) | 28 (21.7%) |  |
| 3 | 34 (26.4%) | 43 (33.3%) |  |
| 4 | 10 (7.8%) | 11 (8.5%) |  |
| 5 | 13 (10.1%) | 8 (6.2%) |  |
| 6 | 4 (3.1%) | 4 (3.1%) |  |
| 7 | 5 (3.9%) | 2 (1.6%) |  |
| 8 | 0 (0.0%) | 2 (1.6%) |  |
| 11 | 1 (0.8%) | 0 (0.0%) |  |

**Table 4.**
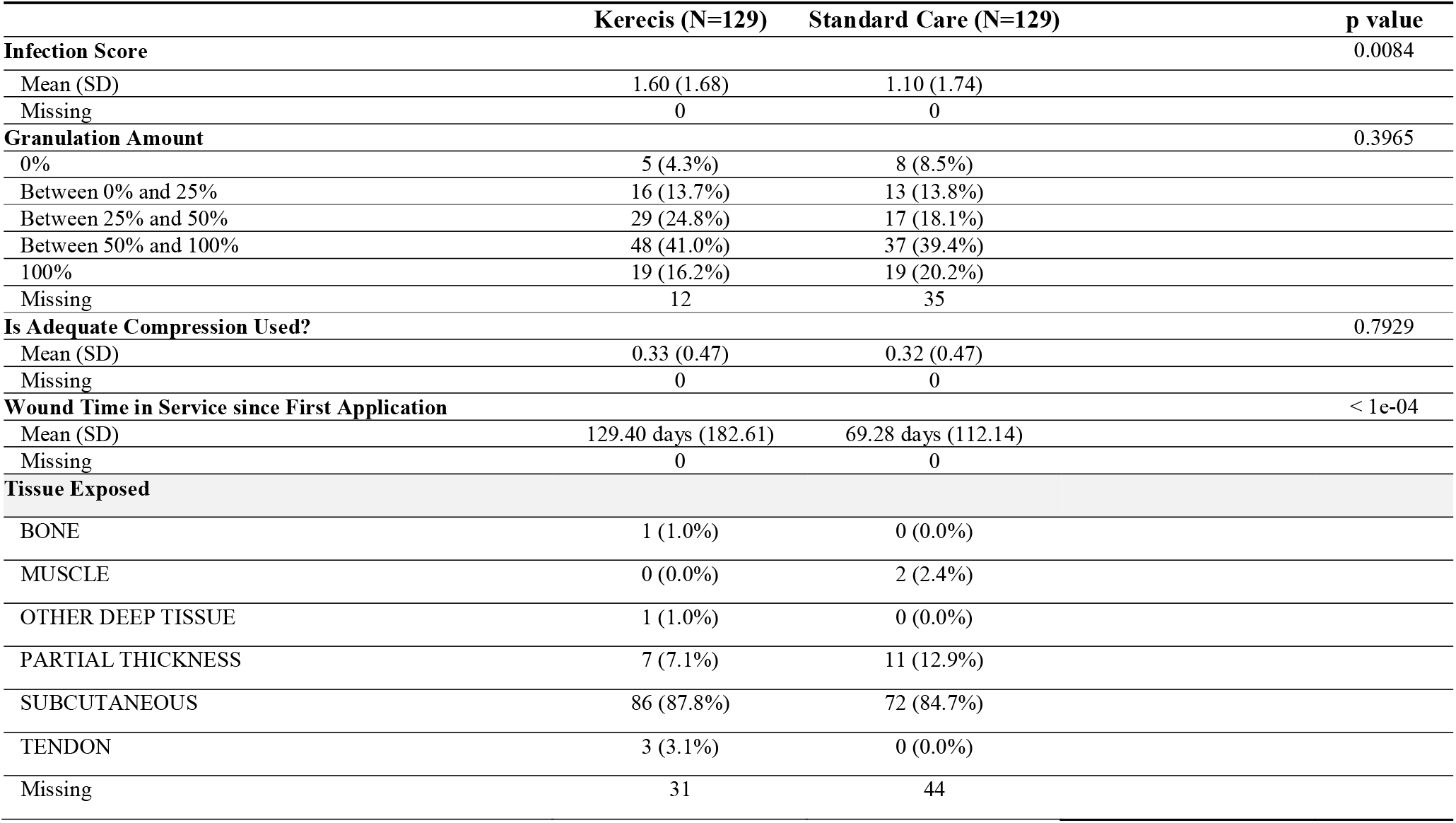
Additional wound baseline and clinical characteristics.

### VLU healing outcome

A total of 85.3% (110/129) of IFSG-treated VLUs healed while 75.2% of SOC-treated VLUs healed (97/129). Although 10.1% more IFSG-treated VLUs healed than SOC-treated VLUs, this difference was slightly below the level of significance (p=0.0801).

### VLU area reduction outcome

While many VLUs decreased in size during treatment, the average reduction in surface area for both IFSG-treated and SOC-treated ulcers was positive, meaning that, on average, ulcers increased in size. On average, IFSG-treated VLUs increased in surface area by 41% (standard deviation of 643%) and SOC-treated VLUs increased in surface area by 62,934% (standard deviation of 615,594%). IFSG-treated VLUs increased in area significantly less than SOC-treated VLUs (p=0.0036).

### Number of IFSG applications

The number of IFSG applications per VLU are given in Table 5. Approximately 59% of VLUs received 3 or fewer IFSG applications, and 79% of patients had 5 or fewer applications. A small percentage (~8.5%) received more than 7 applications.

**Table 5.**

| Number of IFSG applications | Number of VLUs |
| --- | --- |
| 1 | 31 |
| 2 | 21 |
| 3 | 24 |
| 4 | 15 |
| 5 | 11 |
| 6 | 8 |
| 7 | 8 |
| 8 | 2 |
| 9 | 3 |
| 10 | 5 |
| 12 | 1 |

## Discussion

This comparative-effectiveness study leverages a specialty wound-care EHR transmitted to an established national registry to evaluate outcomes for venous leg ulcers under routine practice. Several features of the design collectively reduce the risk of bias to an uncommon degree for retrospective studies and align with best-practice recommendations for real-world evidence. First, cohort construction followed a prespecified plan aligning time-zero between arms at the moment clinical status was identical. This minimizes immortal-time and alignment bias and ensures that baseline covariates reflect the state of the wound and patient at a comparable point in the care pathway. Second, we implemented cohort matching on a comprehensive set of patient- and wound-level variables known to influence healing—including mobility, wound age and size, anatomic location, and the burden of concomitant wounds—followed by formal balance diagnostics using standardized mean differences. Balance was excellent across matched variables, with only small, clinically trivial residual differences. However, wounds that reduce in size by greater than 40% in 4 weeks do not qualify for CTP applications. Therefore, wounds that were more likely to heal are by default potentially enriched in the SOC arm.

Healing rates calculated from RCTs are typically much higher than calculated from real-world data.^13^ One main reason for this phenomenon is that clinical trials typically exclude the patients with the worst prognosis in an effort to obtain patients with a reasonable chance of healing within the timeframe of the study.^13,14,18^ As a result, clinical trials often exclude patients with larger wounds, more severe wounds, and certain comorbidities, creating a mismatch between clinical trial participant populations and real-world patient populations.^13,14,18^ Clinical trials involving VLUs are no exception. For example, one RCT of VLUs drew its participants from a patient population with a 14.1% prevalence of peripheral artery disease (PAD) or atherosclerosis and a 15.5% prevalence of congestive heart failure, but patients with these comorbidities were excluded from the trial.^18^ Further, the mean wound surface area was 31.3 cm^2^ in this patient population, but much lower at only 6.1 cm^2^ in the RCT. Further, as much as 35.9% of the overall patient population had VLUs ≥20 cm^2^, yet the trial did not have any VLUs of this size.^18^ Worryingly, one study showed that using the inclusion/exclusion criteria for 15/17 RCTs involving chronic wounds would result in the exclusion of more than 50% of a patient population drawn from the real world.^14^ However, some recent RCTs have tried to include often overlooked wound types to better reflect the wounds of real world populations. For instance, a recent clinical trial investigating intact fish-skin graft versus standard of care for the treatment of diabetic foot ulcer included wounds of Stage 3 and 4 ulcers, stages often excluded, and included wounds of any size.^19^

In this study, we do not exclude patients or wounds for having any characteristics that might affect tendency to heal. We do not have maximum wound size or age thresholds. We do not exclude any patients based on comorbidities. As a result, we are analyzing a much sicker patient population with more complex wounds than RCT patient populations. The wound outcomes from this study are thus closer to what is seen in the real world. Unexpectedly for such a sick population with complex wounds, healing rates were high for both IFSG-treated and SOC-treated VLUs. At 85.3%, IFSG-treated VLUs had a higher healing rate than the 75.2% of SOC-treated VLUs. This difference was just under statistical significance (p=0.0801), so a larger sample size is needed to determine if treatment with IFSG truly improves healing rate. However, IFSG-treated wounds were statistically significantly older and trended toward being larger than SOC-treated wounds. The fact that IFSG is considered advanced wound care may have created a bias in the IFSG-treated group toward more complex and difficult-to-treat wounds. Further, by excluding SOC-treated wounds historically treated with a skin substitute, we may have biased the SOC-treated group toward less complex and more manageable wounds. Indeed, as much as 21.05% of the IFSG-treated wounds were found to have been treated with another skin substitute historically. The increase in the IFSG-treated wound healing rate just under statistical significance compounded with the bias toward harder-to-manage wounds in the IFSG-treated group strongly suggests that IFSG is effective in improving healing rates in the real-world patient population.

When surface area reduction was calculated, it was found that both IFSG-treated VLUs and SOC-treated VLUs had positive average area reductions, meaning that, on average, VLUs became larger over time. On average, IFSG-treated VLUs had a smaller increase in area than SOC-treated VLUs, which reached statistical significance. Thus, IFSG was more effective than SOC in promoting surface area reduction and minimizing surface area expansion.

Fifty-nine percent of patients received 3 or fewer IFSG applications, indicating that few applications are needed for most patients. The vast majority of patients had 5 or fewer applications of IFSG (~79%), although one patient had as many as 12 applications.

## Conclusions

In this comparative-effectiveness study, IFSG was associated with favorable venous-leg-ulcer (VLU) outcomes under routine practice using a low-bias design (comprehensive covariate control, structured outcomes, and prespecified analyses). Strong absolute healing in the standard-of-care (SOC) arm is best explained by baseline advantages inherent to contemporary practice, shorter duration, smaller area, and “never-advanced-therapy” selection, rather than analytic bias, consistent with our prespecified statistical analysis plan. To refine causal inference, a larger study should apply a symmetrical simulated run-in of failed healing (e.g., ≥4 weeks of guideline-concordant SOC without prespecified improvement) to both IFSG and SOC comparators, better aligning the SOC cohort with advanced-therapy eligibility. Scaling sample size while retaining the same bias-control features will increase precision, enable subgroup analyses, and yield more decision-useful estimates for Medicare coverage determinations.

## Data Availability

Data produced in the present study may be available upon reasonable request to the authors.

## Author Contributions

Conceptualization, C.F., H.M.; methodology, C.F., H.M., B.L.; writing—original draft preparation, C.F., H.M., J.L; writing—review and editing, C.F., H.M., J.L. All authors have read and agreed to the published version of the manuscript.

## Institutional Review Board Statement

The planned study will be conducted in accordance with the Declaration of Helsinki and, as a retrospective analysis of deidentified data, is exempt from Institutional Review Board review, which was also confirmed by the Woodlands IRB after study plan review.

## Informed Consent Statement

As a retrospective study with deidentified data, no informed consent was necessary.

## Data Availability Statement

TFhe data is proprietary but is available upon request to the corresponding author.

## Declaration of Generative AI

During the preparation of this work the authors used OpenAI to support drafting and language refinement of portions of this manuscript, including improving clarity, grammar, and flow. The AI tool was not used for data generation, data analysis, interpretation of results, or drawing scientific conclusions. After using this tool/service, the authors reviewed and edited the content as needed and take full responsibility for the content of the publication.

